# COVID-19-associated acute renal failure in critically ill patients correlates with microthrombosis and renal loss of thrombomodulin

**DOI:** 10.1101/2024.03.18.24304157

**Authors:** Matilda Koskinen, Elisabet Englund, Gül Gizem Korkut, Angelina Schwarz, Marie Jeansson

**Affiliations:** Department of Medicine Huddinge, Karolinska Institutet, Huddinge, Sweden; Division of Pathology, Department of Clinical Sciences, Lund University, Lund, Sweden; Department of Laboratory Medicine, Karolinska Institutet, Sweden; Department of Clinical Sciences, Karolinska Institutet, Sweden

**Author notes:** **Corresponding author:** Marie Jeansson, Karolinska Institutet, Department of Medicine, Blickagången 16, Huddinge, Sweden.

**Keywords:** COVID-19, acute kidney injury, thrombosis, angiopoietin-2, thrombomodulin

## Abstract

Critically ill COVID-19 patients have a high degree of acute kidney injury which develops in up to 85% of patients. We have previously shown that circulating levels of angiopoietin-2 increased in critically ill COVID-19 patients correlated to kidney injury, coagulopathy, and mortality. Furthermore, our experiments showed a causal effect on coagulopathy from angiopoietin-2 binding and inhibition of thrombomodulin mediated anticoagulation. In the current study we hypothesize that renal microthrombi may be a mechanism for reduced renal function in critically ill COVID-19 patients, and that local dysregulation of thrombomodulin and angiopoietin-2 may be involved.

To investigate our hypothesis, we utilized postmortem kidney tissue from seven COVID-19 patients treated at the intensive care unit. We evaluated kidney function, thrombosis, tubular injury, fibrosis, glomerulosclerosis, glomerular size as well as renal expression of thrombomodulin and angiopoietin-2. Proximity ligation assay was utilized to evaluate the presence of angiopoietin-2 binding to thrombomodulin. Normal kidney tissue came from the healthy part of six nephrectomies due to cancer.

Our experiments show renal thrombosis in 6/7 COVID-19 patients, on average 14.7 (6.9-22.5) thrombi per mm2. Most COVID-19 kidneys had extensive kidney injury, especially tubular necrosis, but also glomerular enlargement, glomerulosclerosis, and tubulointerstitial fibrosis which in some cases most likely resulted from underlying disease. Thrombomodulin expression was reduced in glomeruli and peritubular capillaries in kidneys from COVID-19 patients, whereas no change was found for angiopoietin-2.

In summary, our study describes a high degree of acute renal failure, renal microthrombosis, and loss of thrombomodulin in postmortem tissue from critically ill COVID-19 patients.

## Introduction

The corona virus SARS-CoV-2 has manifested as everything from an unnoticeable infection to severe Coronavirus disease-19 (COVID-19) requiring intensive care. While the respiratory system is the main target, the infection is characterized by a broad spectrum of clinical manifestations denoting a multi-organ disease, including systemic inflammation, vascular dysfunction, coagulopathy, hypoperfusion, and multi-organ failure.

Up to 85% of critically ill COVID-19 patients develop acute kidney injury (AKI) (1-4). AKI is a serious syndrome of wide spectrum ranging from functional impairment to renal replacement therapy (5). COVID-19 has both direct and indirect effects that can lead to AKI. Potential direct effects on the kidney include endothelial damage from viral entry, complement activation, and local inflammation (6). Indirect mechanisms for AKI may be more common and include hypercoagulability and thromboembolic disease, systemic inflammation, sepsis, and the use of nephrotoxic medications (6). Fibrin occlusions and thrombi located in glomerular and peritubular capillaries has been found in several studies (7-10), but there are also reports of lack of thrombi in kidney tissue from COVID-19 patients (11). It is well known that AKI predisposes for serious future complications including chronic kidney disease also in COVID-19 patients (12), hence gaining mechanistic insight for AKI in critically ill COVID-19 patients is extremely important.

We recently described a causal effect on hypercoagulability by the inflammatory cytokine angiopoietin-2 (ANGPT2) in vivo (13). ANGPT2 can bind and inhibit thrombomodulin (THBD) mediated protein C activation and anticoagulation (13, 14). THBD is constitutively expressed on the luminal surface of endothelial cells and is an important member of the intrinsic anticoagulation pathway (15, 16). Notably, injection of ANGPT2 in mice resulted in shedding of thrombomodulin from the endothelial surface, hence increased circulating THBD (13). Circulating levels of both ANGPT2 and thrombomodulin are increased in critically ill COVID-19 patients where it correlated with mortality (13, 17-21). Furthermore, we showed that ANGPT2 levels correlated to hypercoagulability and reduced renal function in COVID-19 patients (13). A more well-known role for ANGPT2 is as an antagonist of endothelial TIE2 signaling. TIE2 is a major player in maintenance of endothelial integrity and vascular protection, normally stimulated by its agonistic ligand angiopoietin-1 (ANGPT1) (22). ANGPT2 antagonism of TIE2 and reduced ANGPT1-TIE2 signaling is known to cause endothelial dysfunction, inflammation, vascular leakage, and increased transcription of ANGPT2 (23-29), and this may also contribute to endothelial dysfunction in COVID-19 patients.

We hypothesize that AKI in critically ill COVID-19 patients could be caused by microthrombi in the kidney. To investigate this, we studied postmortem kidney tissue from COVID-19 patients (n=7) treated in the intensive care, which we presumed from our previous study had the highest levels of circulating ANGPT2 (13). Normal renal tissue came from the healthy part of the kidney from patients that underwent complete nephrectomy for kidney cancer (n=6). Kidney tissue was investigated with regards to microthrombi, tubular necrosis, tubulointerstitial fibrosis, glomerulosclerosis, and expression of ANGPT2 and THBD.

## Materials and Methods

### Study design and patients

The present study analyzed postmortem renal tissue and clinical data from critically ill patients with COVID-19, approved by the national Ethical Review Agency (EPM; No. 2021-03959 and 2022-03936-02). Informed consent was obtained from the patient, or next of kin if the patient was unable to give consent. The Declaration of Helsinki and its subsequent revisions were followed. STROBE guidelines were followed for reporting clinical data. All patients died in 2020 or 2021. Samples came from the Region Syd Biobank with the inclusion criteria of COVID-19 by positive reverse-transcription PCR from nasopharyngeal swabs and care at the intensive care unit. AKI was not an inclusion criterion. In total, samples from 7 patients were included in the study. Patients were numbered 1-7, this number is not connected to any patient information known to anyone outside of the research group. Time to autopsy after death in cold storage was 4-7 days. From 2 patients, C19-2 and C19-5, we had one extra sample to investigate if there were large variations within the same patient. These duplicates showed very similar data and were included in the average from each patient. Normal human renal tissue came from visually unaffected part of the kidney from patients that underwent complete nephrectomy for kidney cancer (n=6). All samples were formalin fixed and paraffin embedded tissues. Clinical parameters from COVID-19 patients were evaluated through the web-based clinical records system Melior and included gender, age, medical history, and clinical chemistry of plasma including CRP, IL6, ferritin, D-dimer, fibrinogen, platelet count, creatinine, eGFR, and troponin T. Clinical data was not available for control kidney individuals.

### Kidney histology and microthrombosis

Kidney sections were stained with Masson Trichrome and for fibrinogen (A0080, Dako) by Uppsala University Hospital, Clinical Pathology, FoUU Service. Fibrinogen staining resulted in a lot of background staining, probably due to necrotic tubular epithelial cells, and were not used for quantification. SARS-CoV-2 virus was stained with a rabbit anti-SARS-CoV-2 nucleoprotein antibody (NB100-56576, Novus Biologicals). In addition, samples were stained with Martius Scarlet Blue (MSB) (Atomic Scientific, RRSK2-200) according to the manufacturer’s instructions. Samples were scanned in a slide scanner (AxioScan, Zeiss) with a x20 objective and analyzed using QuPath 4.0 software. The number of thrombi and sclerotic glomeruli was counted manually in each sample and expressed as thrombi/mm^2^ and sclerotic glomeruli/total glomeruli, respectively. In total >3000 glomeruli were investigated, and tissue size ranged between 110-214 mm^2^ in COVID-19 kidneys and 33-94 mm^2^ in control samples. Glomerular size was measured on all samples. Tubulointerstitial fibrosis was evaluated on Masson Trichrome stained sections and expressed as percent fibrosis/total area.

### Immunohistochemistry for nuclei, angiopoietin-2, thrombomodulin, and Pecam1

Immunofluorescence staining was performed with commercially available antibodies on 10 micrometers thick paraffin sample sections after the following preparations: rehydration with Xylene and an alcohol series (100%, 70%, 50%) and heat mediated antigen retrieval (DAKO, 2369) at pH 6 in 95°C for 20 minutes. Samples were blocked with serum free blocking solution (X0909, DAKO) containing 0.25% Triton X100 (Merck, T9284) for 1 hour at room temperature. The samples were then incubated with primary antibodies diluted 1:200 overnight at 4° C. The following antibodies were used; goat anti angiopoietin-2 (R&D Systems, AF623), rabbit anti thrombomodulin (Invitrogen, PA5-120883), and mouse anti PECAM1 (Abcam, Ab949850D). After washing, sections were incubated for 2h at room temperature with fluorescently conjugated secondary antibodies diluted 1:200. The secondary antibodies were: donkey anti rabbit IgG (Thermo Fisher Scientific, A10043 or A31572), donkey anti goat IgG (Thermo Fisher Scientific, A21432), donkey-anti-mouse IgG (Thermo Fisher Scientific, A10038 or A21202). Cell nuclei were stained with Hoechst (Life Technologies, H3570). After washing, samples were mounted with Prolong Gold mounting media (P36930, Thermo Fisher Scientific).

Samples were imaged using a Leica SP8 confocal microscope at x400 magnification. Images were taken of the renal cortex of both the tubular compartment and of PECAM1 positive glomeruli (to exclude sclerotic glomeruli). Images were analyzed with ImageJ (NIH) and area of staining for nuclei ANGPT2, THBD were expressed as the percentage of the entire image area for each image. Images from glomeruli (5-15) and cortex (5-15) for each patient were averaged and used for group comparisons. The nuclei staining area was used as a measurement of tubular health, as necrotic tubular epithelial cells lost nuclear staining.

### Proximity ligation assay

Proximity ligation assay labels antibodies in close proximity (<40 nm) and was utilized to investigate the interaction between ANGPT2 and THBD. The tissue was prepared as previously described using the same primary antibodies. ANGPT2/THBD-complexes were detected with the DuoLink system (DUO02021, Sigma-Aldrich) and DuoLink in Situ Detection Reagents Red (DUO92008, Sigma-Aldrich) according to the manufacturer’s instructions. Secondary antibodies were donkey anti rabbit IgG PLUS (DUO092002, Merck) and donkey anti goat IgG MINUS (DUO92006, Merck). Mounting and imaging procedures were performed as described above.

### Statistics

Data are presented as mean (SD) in graphs and mean (95% CI) in tables. Means between groups were compared using unpaired two-tailed Student’s t-test (for 2 groups) utilizing GraphPad Prism version 10 (GraphPad Software Inc.). Due to uneven distribution in many groups, the data was natural log-transformed before statistical analysis as previously suggested.(30) A *P*<0.05 was considered statistically significant.

## Results and Discussion

### General and Clinical Characteristics

The demographic and clinical features of the study participants are shown in Table 1. The critically ill COVID-19 patients, comprising two females and five males included in the study died 4-26 days after hospitalization (mean 11 days). At time of death, their mean age was 69 (range, 59-75). Five subjects had various comorbidities of varying severity, including hypertension, diabetes, chronic kidney disease, pulmonary diseases, and a history of thromboembolic events. None of them had severe heart failure. Two subjects had no preexisting comorbidities at the onset of COVID-19 infection. One individual had sepsis prior to death and all patients were treated in an intensive care unit. Four patients received invasive ventilation and one patient extracorporeal membrane oxygenation (ECMO) (Table 1). Clinical characteristics were not available from whom the control kidneys tissue originated.

**Table 1.**
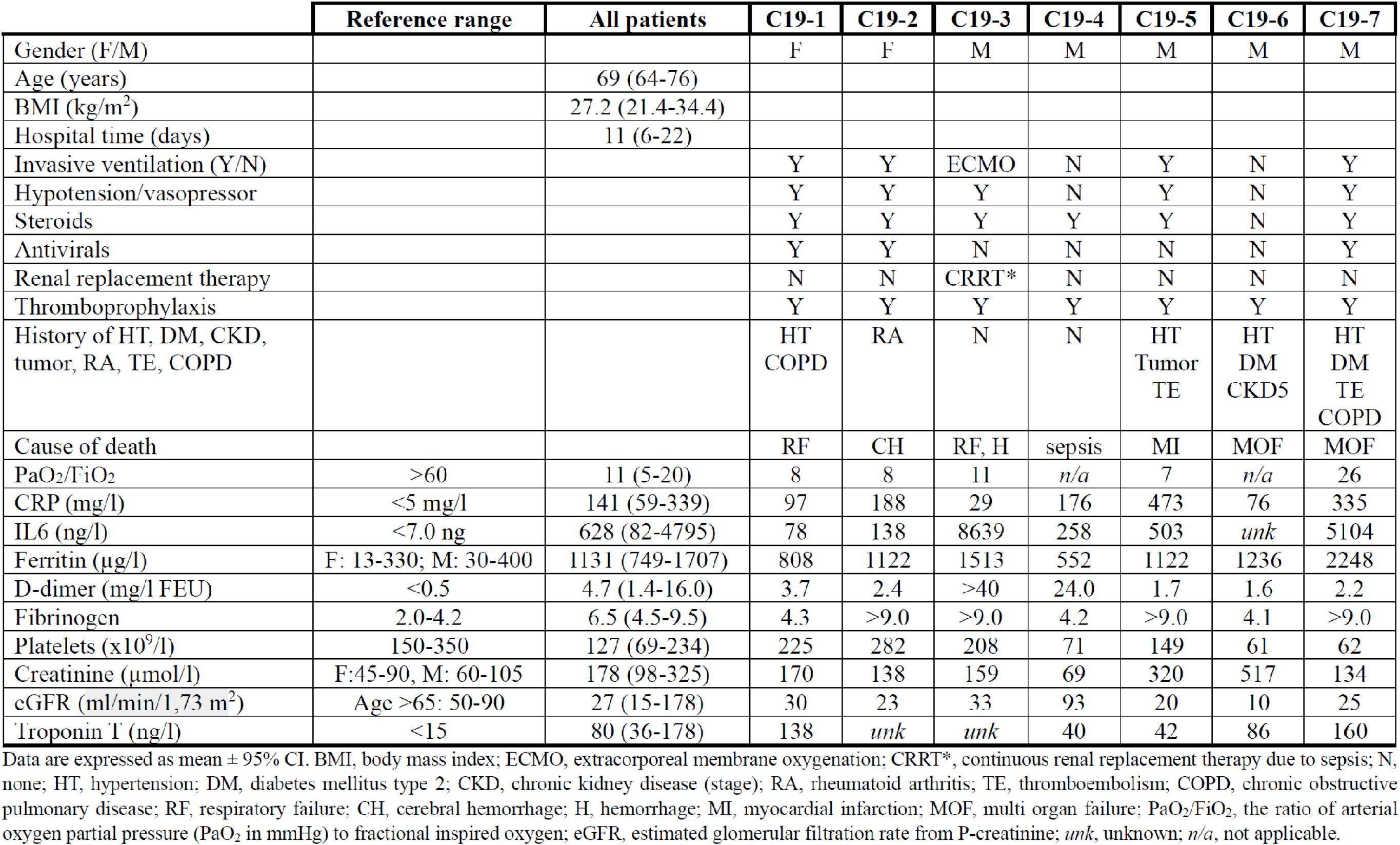
Patient demographics and clinical parameters.

### Acute kidney injury (AKI) in a majority of critically ill COVID-19 patients

Many studies have reported a high incidence of AKI in COVID-19 patients (1-4). Reports suggest that COVID-19 may affect the kidney by direct virus-mediated injury, cytokine storm, dysregulation of complement, and hypercoagulability (31). AKI associates with higher mortality, increased organ failure, and prolonged intensive care stay (4). In the current study, 6/7 COVID-19 patients had AKI during their hospital stay as seen by both increased P-creatinine and decreased estimated glomerular filtration rate (eGFR) (Table 1). One patient (C19-4) had normal kidney function and no history of comorbidities before COVID-19. One patient (C19-6) had chronic kidney disease stage 5 (CKD5) with an eGFR of 10 ml/min/1.73 m^2^. None of the other patients had a history of kidney injury.

In the current study, renal tubular necrosis was extensive in COVID-19 kidneys in agreement with numerous reports (7,8,27,28). To estimate tubular health, we quantified the nuclear area labelled with Hoechst staining from confocal images of renal cortex without glomeruli. Control kidneys had a nuclear area of 9.8% which was significantly reduced to 0.9% in COVID-19 kidneys (*P*<0.001; Fig. 1A, B; Table 2). It cannot be ruled out that some autolysis occurred of tubular epithelial cells, however, the presence of viable tissue suggests that this is not a major contributor to necrotic cells.

**Table 2.**
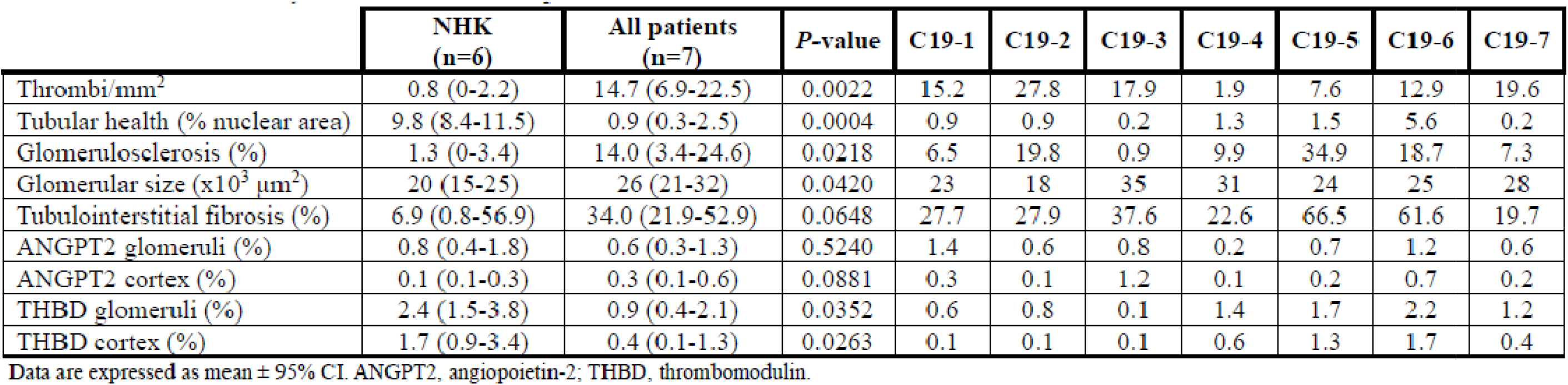
Postmortem kidney data from COVID-19 patients and controls.

**Figure 1.**
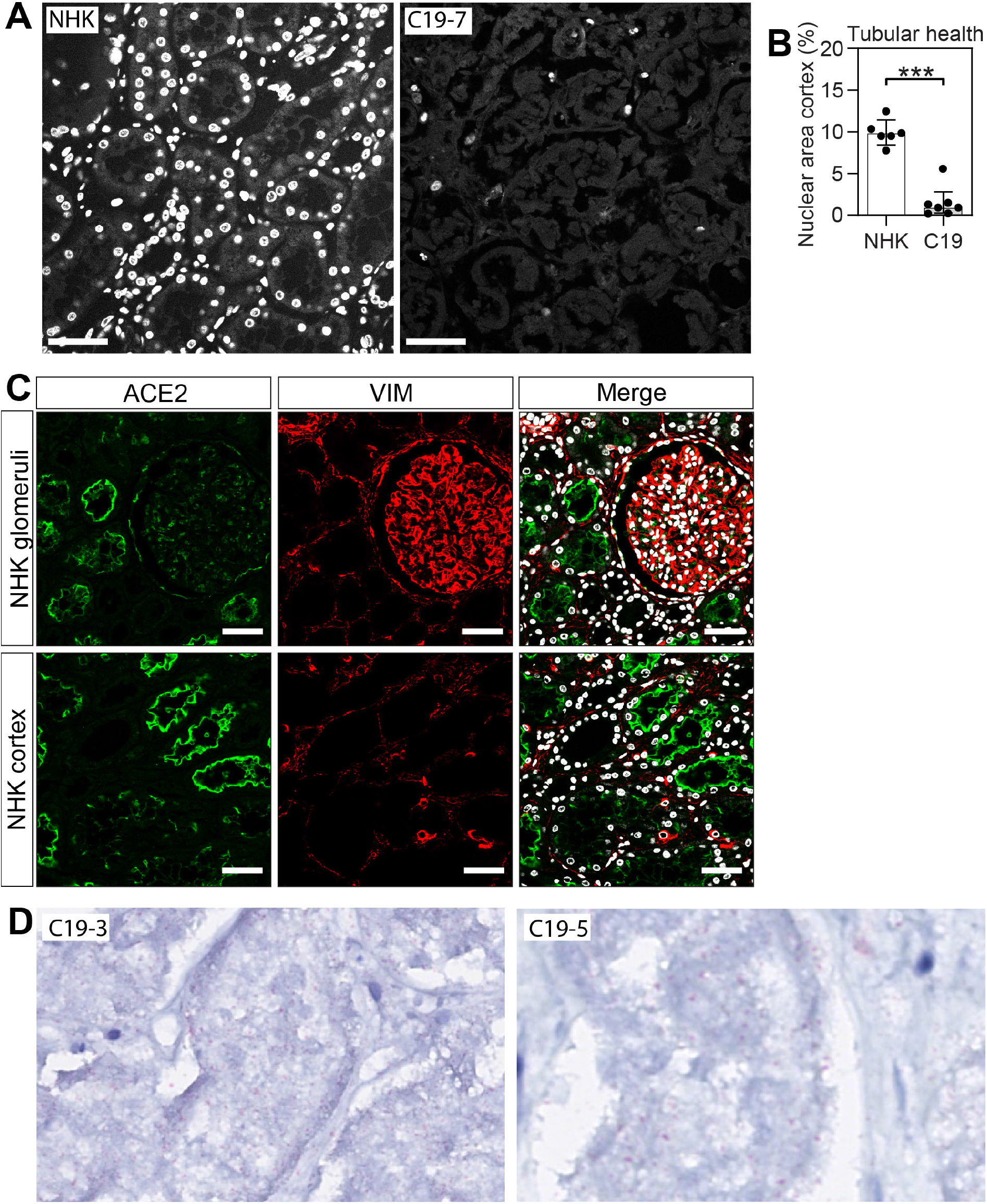
Tubular health and ACE2 expression in human kidney. (**A**) The nuclear staining by Hoechst in fluorescence immunohistochemistry of renal cortex samples from normal human kidneys (NHK, n=6) and kidneys from COVID-19 patients (C19, n=7). (**B**) The area of nuclear staining was quantified and used as a measure of tubular health. (**C**) Fluorescence immunohistochemistry of ACE2 (green) expression in proximal epithelial cells and vimentin (VIM, red) in podocytes and mesenchymal cells in NHK. Original magnification x400, scale bars = 50 μm. (**D**) SARS-CoV-2 virus staining (red chromophore) in tubular segments. Data are presented as mean (SD). \*\*\**P*<0.001 with statistical analysis between groups using Student’s t test (unpaired, 2-tailed).

One proposed mechanism for the extensive tubular necrosis seen in critically ill COVID-19 patients is viral entry into tubular epithelial (40), via angiotensin-converting enzyme 2 (ACE2) receptor. Our staining of ACE2 showed extensive expression in tubular segments in control kidneys (Fig. 1C), in agreement with other studies (40-42). SARS-CoV-2 virus was evident in COVID-19 patients, especially in tubular segments (Fig. 1D).

### Renal thrombosis is common and associated with COVID-19-AKI

Renal thrombosis was evaluated with Martius Scarlet Blue staining on postmortem kidney tissue. Thrombi were found in both glomerular and peritubular capillaries with an average of 14.7 thrombi/mm^2^ in COVID-19 kidney tissue compared to 0.8 thrombi/mm^2^ in control kidneys (*P*<0.01) (Fig. 2; Table 2). It should be noted that one COVID-19 patient (C19-4) only had 1.9 thrombi/mm^2^, which is similar to some controls. Importantly, this is the only patient without signs of AKI, as seen by the normal range of P-creatinine (69 μmol/l) and eGFR (93 ml/min/1.73m^2^) (Table 1). Our study is to our knowledge the first attempt at quantification of renal microthrombi in individual patients. This was in part possible due to the rather large tissue samples we obtained for the study (see Methods). Other studies have reported an incidence of renal microthrombi in 5/6 COVID-19 patients (7) and in 6/8 COVID-19 patients (9). Three other studies have reported incidences of microthrombi in the range of 4.5-14% (32-34). The patient records of clinical data showed that all COVID-19 patients in the current study had increased D-dimer levels, and either borderline high or increased fibrinogen (Table 1), in line with a hypercoagulative state. All patients received thromboprophylaxis. Although two patients had a history of thromboembolism, none of the patients were diagnosed with pulmonary embolism during their hospital stay despite investigation.

**Figure 2.**
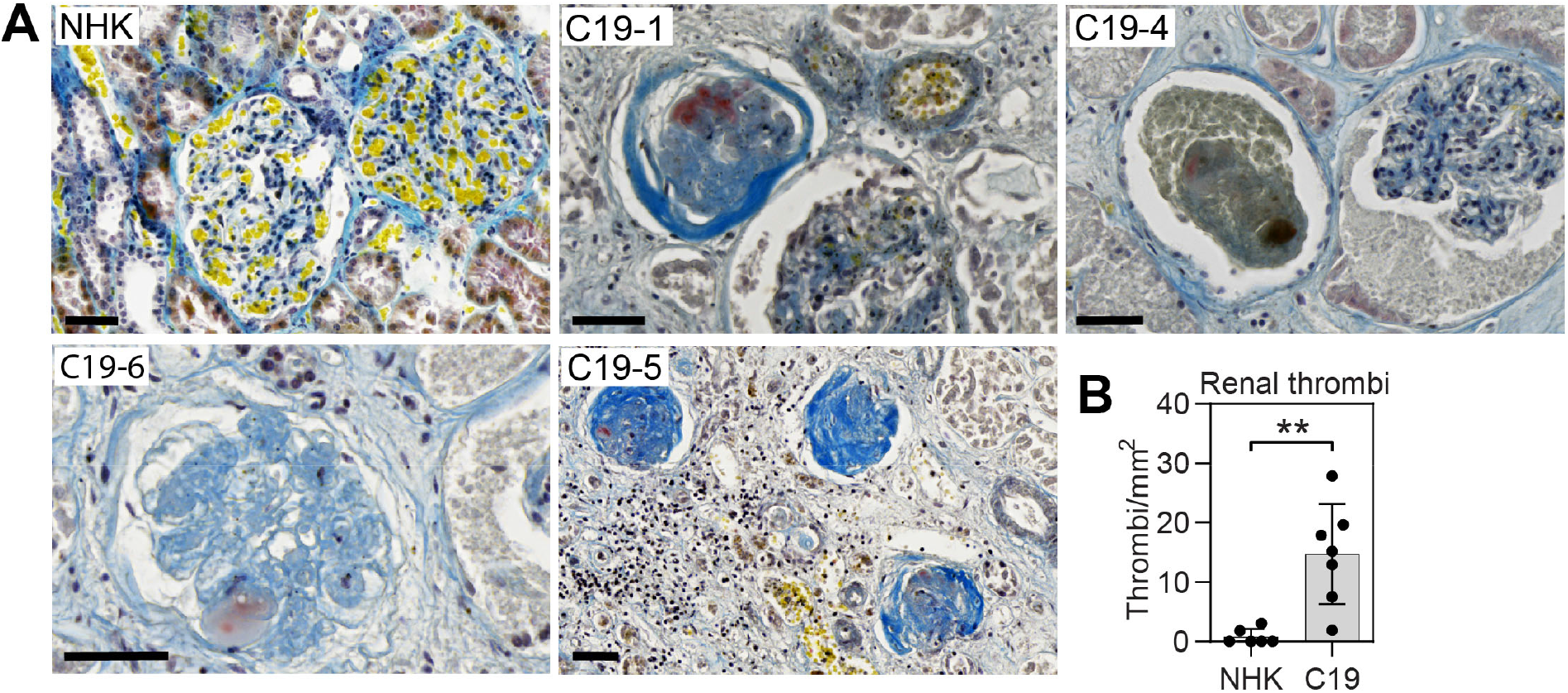
Renal thrombosis and tubular necrosis in critically ill COVID-19 patients. (**A**) Martius Scarlet Blue (MSB) staining of kidney tissue from critically ill COVID-19 patients (C19) and normal human kidney (NHK). MSB stains fibrin (red), fresh fibrin (yellow), erythrocytes (yellow), and connective tissue (blue). Original magnification x200, scale bars = 50 μm. (**B**) Quantification of thrombi from MSB stained tissue expressed as thrombi/mm^2^, *n*=6 (NHK) and *n*=7 (C19). Original magnification x400, scale bars = 50 μm. Data are presented as mean (SD). \*\**P*<0.01 with statistical analysis between groups using Student’s t test (unpaired, 2-tailed).

### Renal THBD decrease suggest endothelial dysfunction and locally reduced anticoagulation

THBD is an important member of the intrinsic anticoagulation pathway and expressed on the luminal surface of endothelial cells (15, 16). Endothelial-specific knockout of THBD in mice disrupts the activation of protein C and causes lethal thrombus formation (35), highlighting the potency of this pathway. Circulating THBD is derived from membrane THBD by endothelial shedding and is a marker of endothelial dysfunction (36, 37). Loss of THBD from lung endothelium has been shown in COVID-19 (19, 38), but to our knowledge no data is available for the kidney in COVID-19. Staining of postmortem kidneys showed a significant loss of THBD in COVID-19 kidneys in both peritubular capillaries and glomerular capillaries compared to controls (Fig. 3; Table 2). In peritubular capillaries THBD expression decreased from 1.7% in control kidneys to 0.4% in COVID-19 kidneys (*P*<0.05). The same pattern was seen in glomerular capillaries with 2.4% THBD in controls compared to 0.9% in COVID-19 kidneys (*P*<0.05).

**Figure 3.**
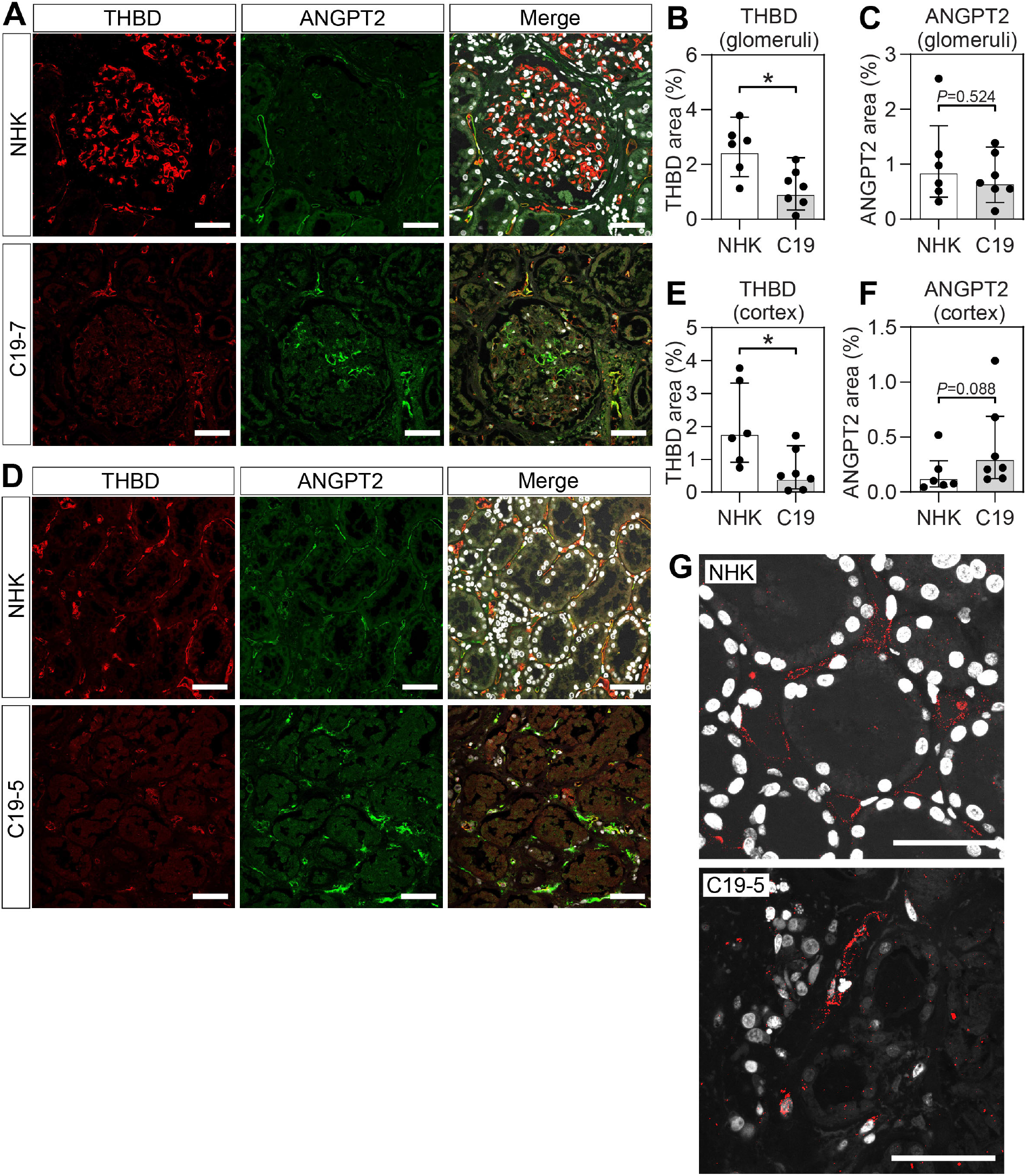
Renal thrombomodulin and angiopoietin-2 expression in critically ill COVID-19 patients. Representative images of immunohistochemistry and quantification of thrombomodulin (THBD) and angiopoietin-2 (ANGPT2) expression in (**A-C**) glomeruli and (**D**-**F**) cortex of normal human kidneys (NHK, *n*=6) and kidneys of COVID-19 (C19, *n*=7) patients. (**G**) Proximity ligation assay shows thrombomodulin and angiopoietin-2 complexes (red) and nuclei (white) in peritubular capillaries of NHK and C19 kidney, respectively. Original magnification x400, scale bars = 50 μm. Data are presented as mean (SD). \**P*<0.05 with statistical analysis between groups using Student’s t test (unpaired, 2-tailed).

We also investigated the renal expression of ANGPT2. In contrast to THBD, we did not find changes in ANGPT2 expression in COVID-19 kidneys compared to controls neither in peritubular capillaries nor in glomerular capillaries (Fig. 3; Table 2). In agreement with this, Volbeda *et al* described a similar *ANGPT2* expression pattern in renal postmortem biopsies from COVID-19 patients with AKI compared to controls (7). Although circulating levels were not available for the patients included in the current study, we would assume ANGPT2 levels are elevated based on published data concerning critically ill COVID-19 patients (13, 17-21). In a previous study, we showed that ANGPT2 injection in mice resulted in rapid increase of circulating THBD, and loss of THBD expression in lung, suggesting shedding from the endothelial surface (39). In summary, local loss of THBD most likely indicates a prothrombotic state, potentially influenced by increased circulating levels of ANGPT2.

We were also interested in visualizing the protein-protein interaction between THBD and ANGPT2. To achieve this, we utilized proximity ligation assay that detects protein-protein interaction in close proximity (<40 nm). The assay showed THBD/ANGPT2 interaction in peritubular capillaries (Fig. 3G). However, although the presence of complexes could be seen, the signal was variable, and no conclusions could be made on differences between COVID-19 kidneys and controls. More studies are needed to investigate the THBD/ANGPT2 interaction more precisely in the context of kidney injury.

### Glomerulosclerosis, glomerular size and tubulointerstitial fibrosis in COVID-19-AKI

Masson trichrome staining of kidneys was utilized to investigate glomerulosclerosis, glomerular size and tubulointerstitial fibrosis in kidneys from COVID-19 patients and controls. All glomeruli (>3000) in the postmortem kidneys were evaluated and glomerulosclerosis were expressed as a percentage of glomerulosclerotic glomeruli/total glomeruli. COVID-19 afflicted kidneys exhibited an average of 14.0% glomerulosclerosis compared to 1.3% observed in control kidneys (*P*<0.05; Fig. 4A, B; Table 2). Patient C19-5 with a glomerulosclerosis rate of 34.9% had no previous diagnosis of kidney disease. Two other patients, C19-2 and C19-5 had high degrees of glomerulosclerosis as well, at 19.8% and 18.7%, respectively. Patient C19-5 had a history of diabetes and CKD5 which can explain the high degree of glomerulosclerosis. Patient C19-2, on the other hand, had rheumatoid arthritis but no previous history of kidney disease. Previous studies have not mentioned glomerulosclerosis as a result of COVID-19, instead it has been linked to underlying chronic kidney disease (7,8,27,28).

**Figure 4.**
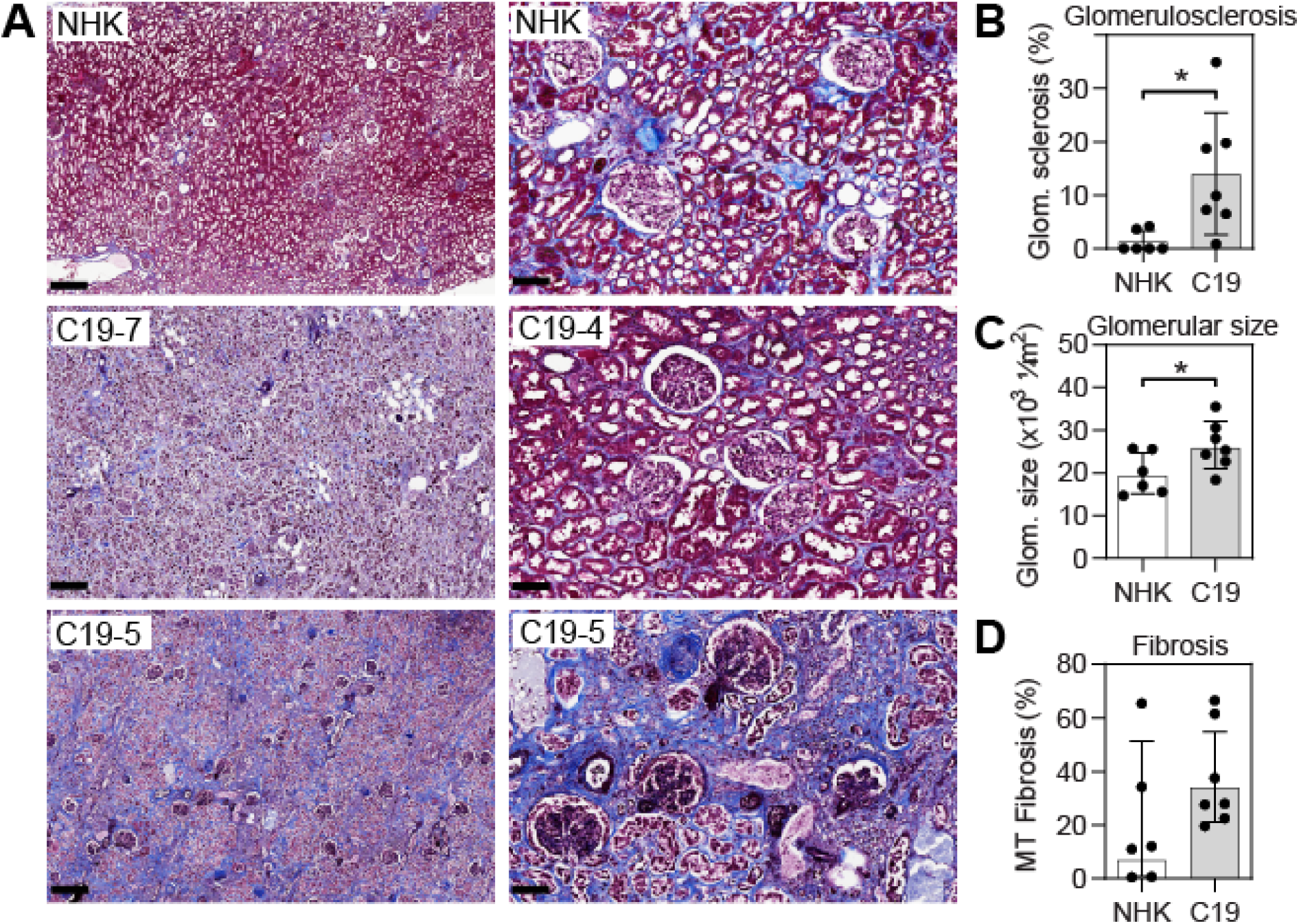
Glomerulosclerosis, glomerular size, and fibrosis in critically ill COVID-19 patients. (**A**) Representative images of Masson trichrome staining of kidneys from normal human kidney (NHK) and COVID-19 patients (C19). Original magnification x200, scale bars = 400 μm (left images) and 100 μm (right images). Quantification of (**B**) glomerulosclerosis and (**C**) glomerular size from MSB stained sections and (**D**) tubulointerstitial fibrosis from Masson trichrome stained sections from NHK (*n*=6) and C19 patients (*n*=7). Data are presented as mean (SD). \**P*<0.05 with statistical analysis between groups using Student’s t test (unpaired, 2-tailed).

During the analysis of glomerulosclerosis, it was noted that some patients had enlarged glomeruli. The glomerular area was then measured and compared between control and COVID-19 kidneys. The average glomerular size in control kidneys were 20×10^3^ μm^2^, consistent with the findings from previous studies (43, 44), whereas kidneys from COVID-19 patients were significantly enlarged with an average size of 26×10^3^ μm^2^ (*P*<0.05; Fig. 2A, C; Table 2). The significant increase in the mean glomerular size was primarily due to enlarged glomeruli in patients C19-3, C19-4, and C19-7. Of these, C19-3 and C19-4 had no history of comorbidities and C19-7 had diabetes type 2, a known cause of enlarged glomeruli (44).

When evaluating tubulointerstitial fibrosis it was evident that two control kidneys also had extensive fibrosis, the reason for this is unknown, but it could be due to old age. The COVID-19 groups showed notable increase in tubulointerstitial fibrosis with patient C19-5 and C19-6 demonstrating particularly high levels at 66.5% and 61.6% respectively. As mentioned, patient C19-5 had CKD5 accounting for the high degree of tubulointerstitial fibrosis. In contrast, patient C19-6 did not have a history of kidney disease.

In conclusion, our study of seven critically ill COVID-19 patients showed substantial tubular necrosis in all cases, with six patients having AKI. The same six individuals also had substantial renal microthrombosis, while other renal pathologies were not uniformly present. Endothelial dysfunction and localized loss of THBD most likely indicate a prothrombotic state, potentially influenced by increased circulating levels of ANGPT2.

## Author contributions

Conceptualization, M.J.; Data curation, M.K. and M. J.; Formal analysis, M.K. and M.J.; Methodology, M.K., and M.J.; Recourses, E.E., G.G.K., A.S., and M.J.; Writing – original draft, M.K. and M.J.; Writing – review and editing, E.E., G.G.K., and A.S. All authors have read and agreed to the published version of the manuscript.

## Institutional Review Board Statement

The study was conducted in accordance with the Declaration of Helsinki and approved by the Swedish National Ethical Review Agency (EPM approval No: 2021-03959 and 2022-03936-02) for studies involving humans. STROBE guidelines were followed for reporting clinical data.

## Informed Consent Statement

Informed consent was obtained from the patient, or next of kin if the patient was unable to give consent.

## Data Availability Statement

The authors declare that all supporting data is available within the article.

## Acknowledgment

We thank Professor Jaakko Patrakka at Karolinska Institutet for assistance with control kidney tissue, and Tommy Josefsson at Region Skåne for compiling medical records from included patients.

